# Clinical Outcomes and Plasma Concentrations of Baloxavir Marboxil and Favipiravir in COVID-19 Patients: An Exploratory Randomized, Controlled Trial

**DOI:** 10.1101/2020.04.29.20085761

**Authors:** Yan Lou, Lin Liu, Hangping Yao, Xingjiang Hu, Junwei Su, Kaijin Xu, Rui Luo, Xi Yang, Lingjuan He, Xiaoyang Lu, Qingwei Zhao, Tingbo Liang, Yunqing Qiu

## Abstract

**Background:** Effective antiviral drugs for COVID-19 are still lacking. This study aims to evaluate the clinical outcomes and plasma concentrations of baloxavir marboxil and favipiravir in COVID-19 patients.

**Methods:** Favipiravir and baloxavir acid were evaluated for their antiviral activity against SARS-CoV-2 in vitro before the trial initiation. We conducted an exploratory trial with 3 arms involving hospitalized adult patients with COVID-19. Patients were randomized assigned in a 1:1:1 ratio into baloxavir marboxil group, favipiravir group, and control group. The primary outcome was the percentage of subjects with viral negative by Day 14 and the time from randomization to clinical improvement. Virus load reduction, blood drug concentration and clinical presentation were also observed. The trial was registered with Chinese Clinical Trial Registry (ChiCTR 2000029544).

**Results:** Baloxavir showed antiviral activity in vitro with the half-maximal effective concentration (EC_50_) of 5.48 µM comparable to arbidol and lopinavir, but favipiravir didn’t demonstrate significant antiviral activity up to 100 µM. Thirty patients were enrolled. The percentage of patients who turned viral negative after 14-day treatment was 70%, 77%, and 100% in the baloxavir, favipiravir, and control group respectively, with the medians of time from randomization to clinical improvement was 14, 14 and 15 days, respectively. One reason for the lack of virological effect and clinical benefits may be due to insufficient concentrations of these drugs relative to their antiviral activities.

**Conclusions:** Our findings do not support that adding either baloxavir or favipiravir under the trial dosages to the existing standard treatment.

## Introduction

In December, 2019, several patients with pneumonia of unknown cause were confirmed to be infected with a novel coronavirus, initially named as 2019-nCoV and now named as SARS-CoV-2, in Wuhan, Hubei province, China. The pneumonia was later named as Coronavirus disease 2019 (COVID-19), and soon drew global attention because of the rapidly increasing patient numbers[1]. As of 17:00 on April 29, 2020, over 3,080,000 cases confirmed in China and other 212 countries. Therefore, the situation is grim for the prevention and control of COVID-19.

Up to now, there is still a lack of effective antiviral drugs for COVID-19. The treatment experience can only draw on the characteristics of other coronaviruses, such as highly pathogenic Severe Acute Respiratory Syndrome Coronavirus (SARS-CoV). The antiviral drugs that have been recommended by Diagnosis and treatment program of the novel coronavirus pneumonia (COVID-19) (Trial Sixth Edition,China) include broad-spectrum antiviral drugs (interferon-α, ribavirin), hemagglutinin inhibitors (arbidol), protease inhibitors (lopinavir/ritonavir), and chloroquine phosphate. The latest data shows that some antiviral drugs, including favipiravir, remdesivir, chloroquine phosphate, have inhibitory effect against SARS-CoV-2 in vitro[2]. However, due to the limited clinical experience of using these drugs in COVID-19 patients, inadequate understanding of their mechanism of action against SARS-CoV-2, the antiviral drugs currently in use need more in-depth research basis in clinical application for COVID-19, and their potential efficacy against the SARS-CoV-2 virus has been controversial.

Baloxavir marboxil and favipiravir are novel inhibitors of the influenza RNA replication process by targeting different protein subunits of the influenza polymerase complex. Briefly, baloxavir marboxil inhibits cap-dependent endonuclease [3], and favipiravir inhibits polymerase basic protein 1[4]. Since the SARS-CoV-2 and the influenza virus are both RNA viruses, baloxavir marboxil and favipiravir are considered to be potentially effective against SARS-CoV-2 by blocking its RNA synthesis. Meanwhile, we found that baloxavir marboxil and favipiravir have antiviral activity against SARS-CoV-2 in vitro. Moreover, favipiravir is a small purine analogue and converted into its active ribofuranosyl 5’-triphosphate metabolite in the cell, which can be incorporated in the growing RNA strand[5]. The antiviral activity of fabiravir in vivo may be stronger than that in vitro. Based on the existing treatment experience and related theoretical basis, we decided to study their clinical efficacy in the treatment of COVID-19.

In this study, in vitro activities of antiviral drugs against SARS-CoV-2 were screened firstly. Then, an exploratory single center, open-label, randomized, controlled trial was conducted to evaluate the efficacy and safety of adding baloxavir marboxil or favipiravir to the current standard antiviral treatment in patients confirmed as COVID-19 who are still positive for the SARS-CoV-2 (ChiCTR2000029544). We also measured the plasma concentrations of these antiviral drugs, compared them to the half-maximal effective concentration (EC_50_) values.

## Methods

### In Vitro Antiviral Assay Against SARS-CoV-2

The antiviral activity was evaluated by quantifying the virus yield in the supernatants of infected cell after treatment by qRT-PCR. For additional detailed operation steps, please refer to Supplementary Material (appendix p1).

### Study Design

This trial was an exploratory single center, open-label, randomized, controlled trial to evaluate the efficacy and safety of adding baloxavir marboxil or favipiravir to the current standard antiviral treatment in patients confirmed as COVID-19 who are still positive for the SARS-CoV-2 (ChiCTR2000029544). Participants confirmed as COVID-19 infection were randomly allocated (1:1:1) to the study after approval by the ethics committees. SAS software was used to generate the random number and the treatment group corresponding to the random number. After the subjects passed screening, the researchers assigned the random number according to the order of enrollment, removed the random envelope according to the random number, and treated the subjects according to the random envelope group and treatment plan. The blind method is not suitable for this trial. Figure 1 shows an overview of this trial. The trial was initiated on February 3, 2020, and the data were collected in The First Affiliated Hospital, Zhejiang University School of Medicine. This study was approved by the Ethics Committee of the First Afliated Hospital, Zhejiang University School of Medicine (2020 llT-7). Specific inclusion and exclusion criteria are listed in Supplementary Material (appendix p2).

**Figure 1.**
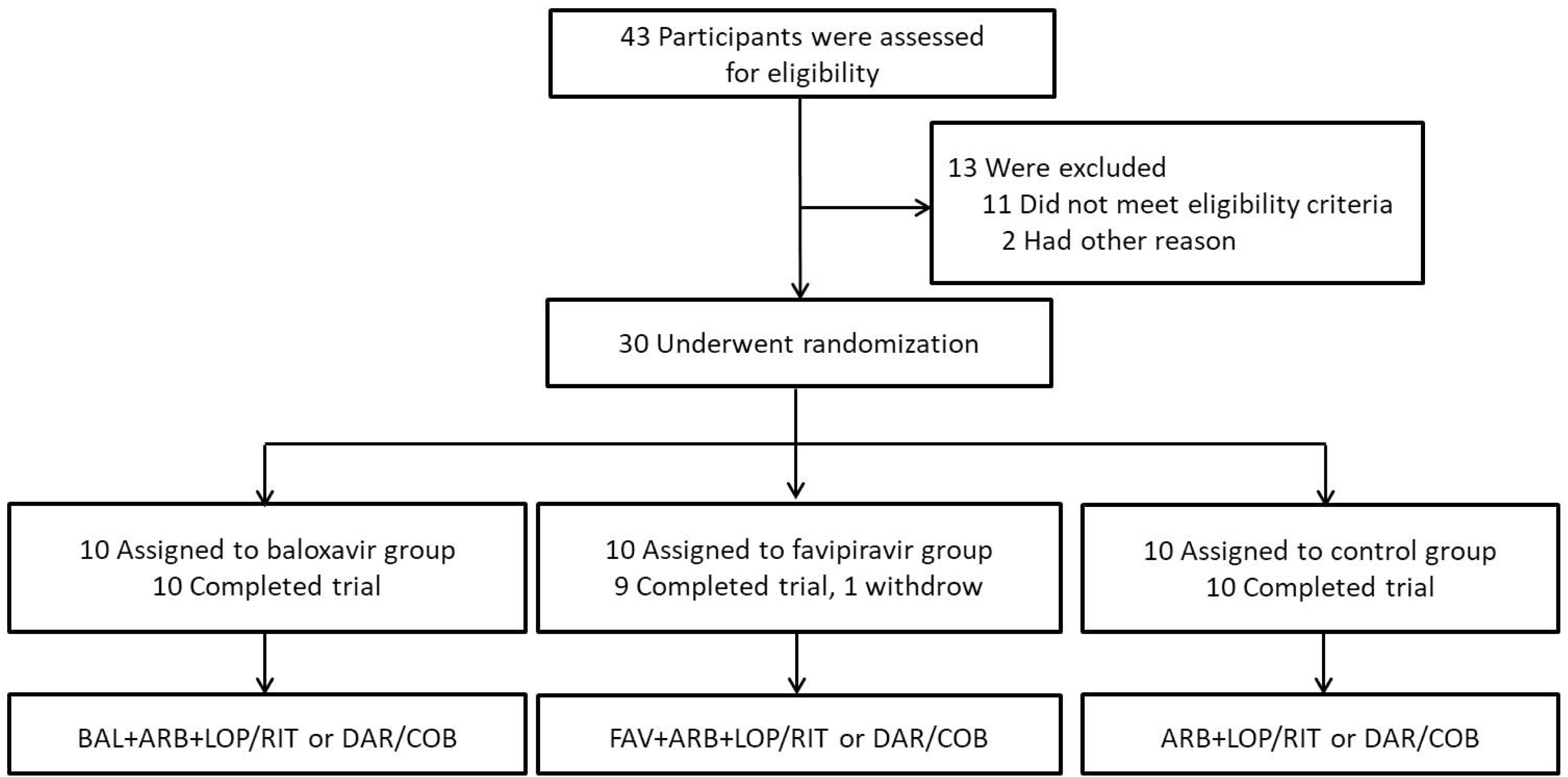
Overview of the Clinical Study. BAL: baloxavir marboxil; FAV: favipiravir; ARB: arbidol; LOP/RIT: lopinavir/ritonavir; DAR/COB: darunavir/cobicistat.

### Dose Administration

All patients start the recommended antiviral treatment (lopinavir/ritonavir, darunavir/cobicistat or arbidol, described below) immediately after the admission diagnosis and received standard care. The trial treatment scheme started as soon as consent was obtained (Day 1). The specific administration is as follows: (1) Baloxavir marboxil group: baloxavir marboxil is used in combination with the existing antiviral treatment (described below). The dose was 80 mg once a day orally on Day 1 and Day 4; for patients who are still positive in virological test, they can be given again on Day 7, no more than three additional doses; (2) Favipiravir group: favipiravir was used in combination with the existing antiviral treatment. The first dose was 1600 mg or 2200mg orally, followed by 600 mg each time, three times a day, and the duration of administration was not more than 14 days; (3) Control group: Continue the existing antiviral treatment.

The existing antiviral treatment included lopinavir/ritonavir (400mg/100mg, bid, po.) or darunavir/cobicistat (800mg/150mg, qd, po.) and arbidol (200mg, tid, po.). All of them were used in combination with interferon-α inhalation (100,000 iu, tid or qid).

## Outcomes

### Primary and Secondary Outcomes

The primary efficacy end point was the percentage of subjects with viral negative by Day 14 and the time from randomization to clinical improvement, defined as the time from randomization to an improvement of two points (from the status at randomization) on a seven-category ordinal scale or live discharge from the hospital, whichever came first. The ordinal scale, which is refer to National Early Warning Score 2 (NEWS2), have been used as end points in clinical trials in patients hospitalized with COVID-19. Secondary clinical end points included the percentage of subjects with viral negative by Day 7, the incidence of mechanical ventilation by Day 14, ICU admission by Day 14, and all-cause mortality by Day14.

### Viral Negativity

SARS-CoV-2 (molecular viral load) was immediately assessed at the hospital laboratory using a semi-quantitative RT-PCR assay (LightCycler 480real-time fluorescent quantitative PCR, Roche). The results are expressed in terms of Ct, whose value is inversely proportional to viral load.

### Drug Concentration Measurement

Blood samples were collected less than one hour after the first dose of favipiravir or baloxavir marboxil on Day 1, then just before dosing on the Day 4 and the Day 7. All samples were immediately shipped to the biosafety level 3 laboratory in our hospital. In this laboratory, the blood samples were heated at 60°C for one hour to inactivate SARS-CoV-2, which did not significantly affect the quantification of these six compounds in plasma (appendix p3). Plasma samples were prepared, frozen at −20°C and transferred to another laboratory in our hospital for drug concentration measurement. Plasma concentrations of baloxavir acid, arbidol, lopinavir, ritonavir and darunavir were determined simultaneously using a validated liquid chromatography-tandem mass spectrometry method which was firstly established in this study (appendix p4). Favipiravir plasma concentrations were analyzed using the liquid chromatography (appendix p4).

### Statistical Analysis

Due to the exploratory nature of this study, a formal sample size calculation would have been of little value and therefore it was not performed. Similarly, more statistical analysis will not be performed.

## Results

### In Vitro Antiviral Activity Against SARS-CoV-2

The activity against SARS-CoV-2 was tested in vitro for the antiviral drugs used in this trial, including arbidol, ritonavir, lopinavir, darunavir, baloxavir acid, and favipiravir. Among the six tested drugs, three drugs showed measurable activity against SARS-CoV-2. Based on the results of non-linear regression fitting, EC_50_ against SARS-CoV-2 was estimated to be 3.32, 5.48, and 10.4 µM, for arbidol, baloxavir acid, and lopinavir, respectively (Figure 2). The antiviral activity of favipiravir was not as effective as observed in a previous study in which an EC_50_ value of 61.88µM was reported[2], and SARS-CoV-2 was inhibited by less than 50% at concentrations up to 100 µM, the highest concentration tested in antiviral assay. Similarly, ritonavir and darunavir did not show antiviral activity against SARS-CoV-2. Furthermore, all the tested drugs had a toxic effect on cells at high concentration but did not show cytotoxicity at the effective concentrations. Together, these results indicate that arbidol, baloxavir acid, and lopinavir could be potential candidates for clinical treatment against COVID-19.

**Figure 2.**
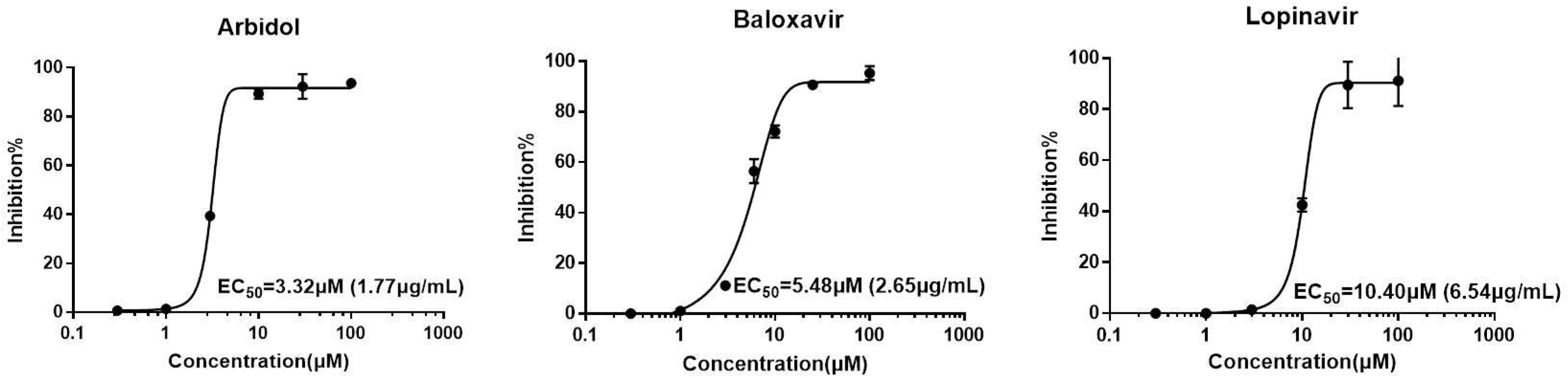
Estimation of in vitro antiviral effect of Arbidol, Baloxavir, and Lopinavir against SARS-CoV-2. Vero E6 Cells were infected with coronovirus strain SARS-CoV-2 in the treatment of different concentrations of the antiviral drugs. The viral yield in the cell supernatant was quantified with qRT-PCR. The Y-axis of the graphs represents inhibition% of virus yield, while the X-axis represents the different concentrations of antiviral drugs in cell culture medium.

### Baseline Characteristics of the Patients

Thirty patients were enrolled in the trial and were randomized into baloxavir marboxil group, favipiravir group, and a control group (Figure 1). One patient in the favipiravir group was subsequently excluded from the final analysis because of discontinuation of favipiravir after Day 1 treatment. The remaining 29 patients were included in the analysis. The baseline characteristics are presented in Table 1. All these patients had no previous history of malignant, chronic obstructive pulmonary disease (COPD), renal insufficiency, and hepatic insufficiency, and there were no deaths during the trial.

**Table 1.** Baseline characteristics of patients with 2019-nCoV infection (COVID-19

| Characteristic | Baloxavir Group | Favipiravir Group | Control Group | Total |
| --- | --- | --- | --- | --- |
| n | 10 | 9 | 10 | 29 |
| Age (year) -mean(SD) | 53.5 (12.5) | 58.0 (8.1) | 46.6 (14.1) | 52.5 (12.5) |
| Male sex - no. (%) | 7 (70) | 7 (77) | 7 (70) | 21 (72.4) |
| Days from symptom onset to randomization-mean(SD) | 12.7 (3.5) | 8.5 (3.7) | 13.6 (4.6) | 11.7 (4.4) |
| Comorbidity- no. (%) |  |  |  |  |
| Diabetes | 0 | 2(22) | 0 | 2 (6.9) |
| Hypertension | 2 (20) | 1 (11) | 3(30) | 6 (20.7) |
| Hyperlipidemia | 0 | 0 | 1 (10) | 1 (3.4) |
| Cardiovascular disease | 3 (30) | 1(11) | 0 | 4 (13.8) |
| NEWS2 score- median (IQR) | 4 (4-5) | 4 (4-5) | 4 (4-5) | 4 (4-5) |
| Ct value- median (IQR) | 28.2 (22.1-36.8) | 25.4 (19.8-36.0) | 29.6 (19.8-37.1) | 28.2 (19.8-37.1) |
| Body temperature (°C) -median (IQR) | 36.9 (36.2-38.4) | 36.9 (36.3-39.6) | 36.9 (36.0-37.9) | 36.9 (36.0-39.6) |
| Fever - no. (%) | 1 (10) | 3 (33) | 2 (20) | 6 (20.7) |
| Respiratory rate >24/min - no. (%) | 0 | 1(11) | 0 | 1 (3.4) |
| Serum biochemistry - median (IQR) |  |  |  |  |
| Haemoglobin (g/dl) | 133 (118-153) | 141 (127-156) | 133 (89-155) | 138 (89-156) |
| Total peripheral WBC count (*10E9/l) | 8.3 (3.3-27.9) | 7.8 (3.9-14.1) | 6.3 (2.9-19.4) | 7.5 (2.9-27.9) |
| Lymphocyte count (*10E9/l) | 0.6 (0.2-2.1) | 0.9 (0.3-1.6) | 0.8 (0.3-1.6) | 0.7 (0.2-2.1) |
| Platelet count (*10E9/l) | 174 (108-459) | 206 (82-281) | 199 (97-347) | (82-459) |
| ALT (U/l) | 22 (10-72) | 21 (8-148) | 27 (12-76) | 22 (8-148) |
| AST (U/l) | 19 (9-44) | 17 (12-77) | 16 (12-44) | 17 (9-77) |
| Albumin (g/l) | 34.8 (28.8-43.6) | 37.8 (29.7-43.0) | 34.9 (31.1-44.6) | 36.8 (28.8-4.6) |
| Creatinine (μmol/l) | 67 (54-83) | 76 (65-104) | 82 (54-91) | 76 (54-104) |
| LDH (U/l) | 265 (219-370) | 252 (174-323) | 307 (142-358) | 249 (142-370) |
| CPK (U/l) | 63 (20-223) | 50 (34-117) | 43 (22-186) | 53 (20-223) |
| CRP (mg/l)* | 14.1 (0.65-50.9) | 27.3 (0.32-79.9) | 2.1 (0.32-26.4) | 10.6 (0.32-79.9) |
| PCT (ng/ml) † | 0.06 (0.03-0.3) | 0.07 (0.02-0.1) | 0.05 (0.02-0.1) | 0.06 (0.02-0.3) |

WBC = white blood cell; ALT = Alanine aminotransferase; AST=Aspartate aminotransferase; LDH = lactate dehydrogenase; CPK=creatine phosphohykinase; CRP=C-reactive protein; PCT=procalcitonin
\*Four missing values (two in Baloxavir group, two in Favipiravir group).
†Five missing values (two in Baloxavir group, two in Favipiravir group, one in Control group).

The demographic characteristics, Ct value, and initial serum biochemistry were balanced in the three groups. Figure 1 shows that although the COVID-19 patients were treated with various existing standard antiviral treatments following the recommendation by the National Health Commission of People’s Republic of China (Trial Sixth Edition), in this trial, the three groups of patients received similar antiviral treatment.

## Clinical Outcomes

### Primary Outcome

A total of 24 (82.8%) patients turned viral negative (defined as two consecutive tests with viral RNA undetectable results) within 14 days after the initiation of the trial. The percentage of patients who turned viral negative after 14-day treatment was 70%, 77%, and 100% in the baloxavir, favipiravir, and control group respectively, of which the control group was higher than that of the other two treatment groups (Figure 3). The medians of time from randomization to clinical improvement were 14, 14 and 15 days in the baloxavir, favipiravir, and control group, respectively (Table 2).

**Figure 3.**
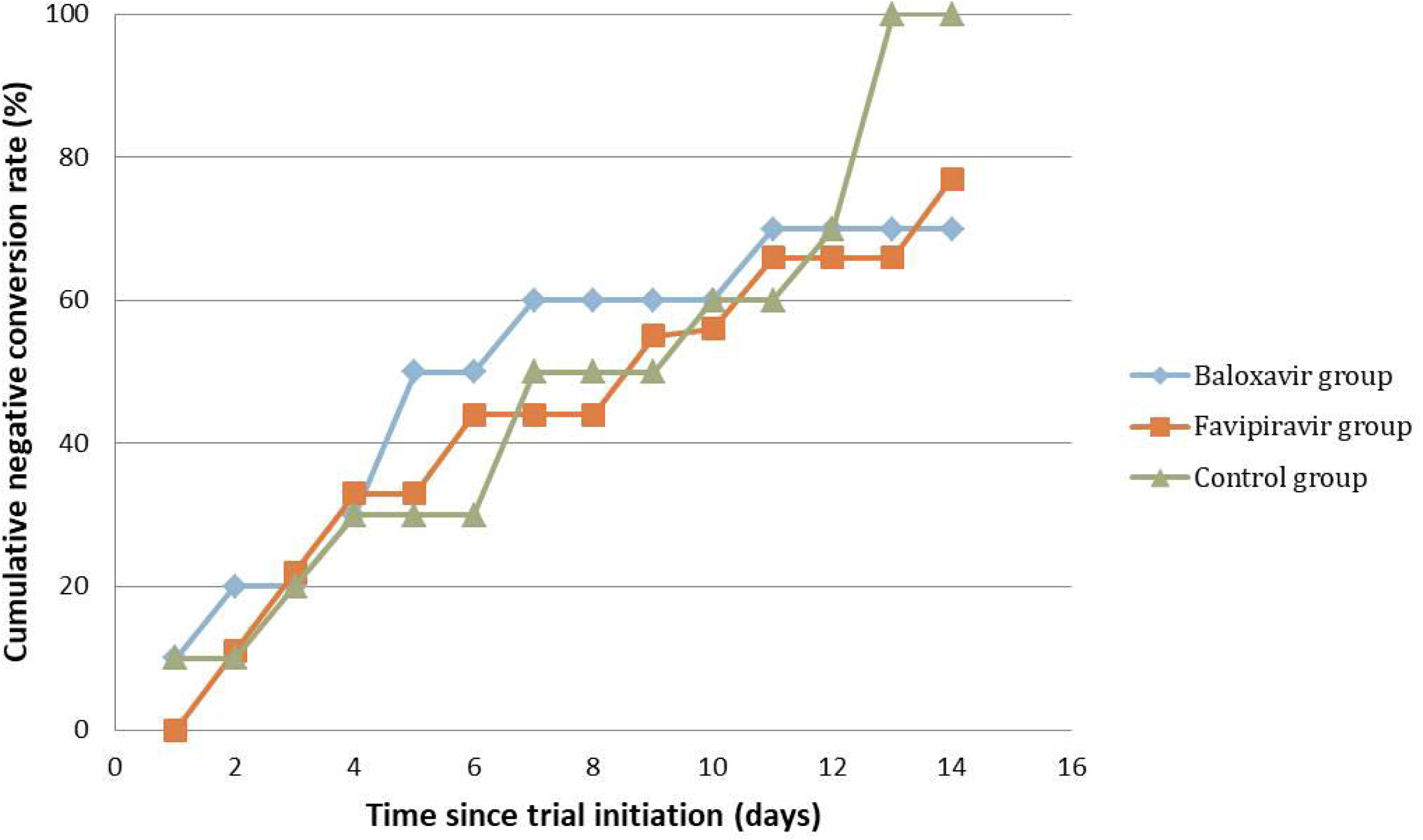
Cumulative negative conversion rates of subjects by Day 14. The x-axis represents the time (days) since trial initiation. The y-axis represents cumulative negative conversion rate.

**Table 2.**
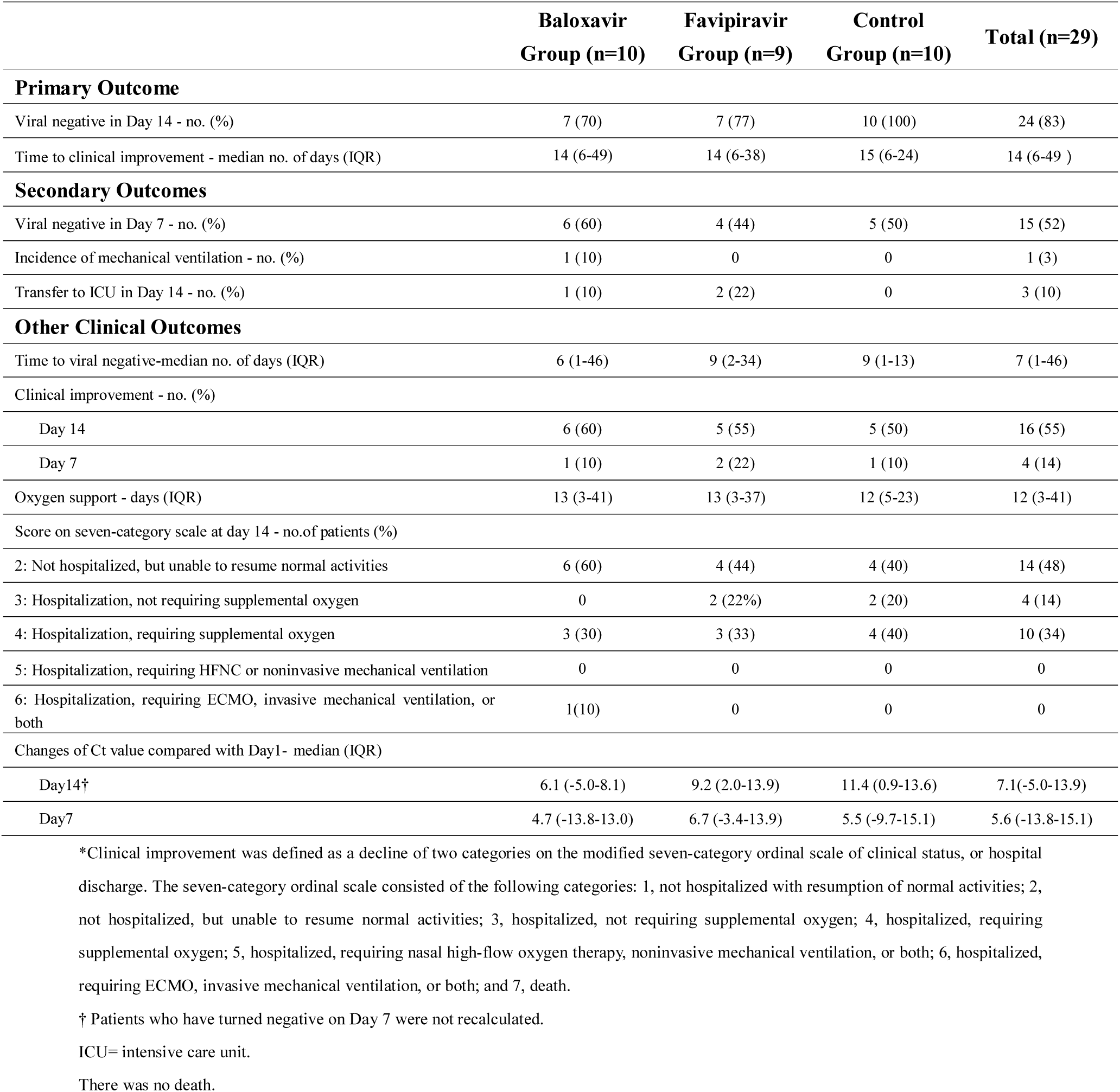
Clinical Outcomes of patients with 2019-nCoV infection (COVID-7)*

|  | **Baloxavir**<br>**Group (n=10)** | **Favipiravir**<br>**Group (n=9)** | **Control**<br>**Group (n=10)** | **Total (n=29)** |
| --- | --- | --- | --- | --- |
| **Primary Outcome** |  |  |  |  |
| Viral negative in Day 14 - no. (%) | 7 (70) | 7 (77) | 10 (100) | 24 (83) |
| Time to clinical improvement - median no. of days (IQR) | 14 (6-49) | 14 (6-38) | 15 (6-24) | 14 (6-49 ) |
| **Secondary Outcomes** |  |  |  |  |
| Viral negative in Day 7 - no. (%) | 6 (60) | 4 (44) | 5 (50) | 15 (52) |
| Incidence of mechanical ventilation - no. (%) | 1 (10) | 0 | 0 | 1 (3) |
| Transfer to ICU in Day 14 - no. (%) | 1 (10) | 2 (22) | 0 | 3 (10) |
| **Other Clinical Outcomes** |  |  |  |  |
| Time to viral negative-median no. of days (IQR) | 6 (1-46) | 9 (2-34) | 9 (1-13) | 7 (1-46) |
| Clinical improvement - no. (%) |  |  |  |  |
| Day 14 | 6 (60) | 5 (55) | 5 (50) | 16 (55) |
| Day 7 | 1 (10) | 2 (22) | 1 (10) | 4 (14) |
| Oxygen support - days (IQR) | 13 (3-41) | 13 (3-37) | 12 (5-23) | 12 (3-41) |
| Score on seven-category scale at day 14 - no.of patients (%) |  |  |  |  |
| 2: Not hospitalized, but unable to resume normal activities | 6 (60) | 4 (44) | 4 (40) | 14 (48) |
| 3: Hospitalization, not requiring supplemental oxygen | 0 | 2 (22%) | 2 (20) | 4 (14) |
| 4: Hospitalization, requiring supplemental oxygen | 3 (30) | 3 (33) | 4 (40) | 10 (34) |
| 5: Hospitalization, requiring HFNC or noninvasive mechanical ventilation | 0 | 0 | 0 | 0 |
| 6: Hospitalization, requiring ECMO, invasive mechanical ventilation, or both | 1(10) | 0 | 0 | 0 |
| Changes of Ct value compared with Day1- median (IQR) |  |  |  |  |
| Day14† | 6.1 (-5.0-8.1) | 9.2 (2.0-13.9) | 11.4 (0.9-13.6) | 7.1(-5.0-13.9) |
| Day7 | 4.7 (-13.8-13.0) | 6.7 (-3.4-13.9) | 5.5 (-9.7-15.1) | 5.6 (-13.8-15.1) |

\*Clinical improvement was defined as a decline of two categories on the modified seven-category ordinal scale of clinical status, or hospital discharge. The seven-category ordinal scale consisted of the following categories: 1, not hospitalized with resumption of normal activities; 2, not hospitalized, but unable to resume normal activities; 3, hospitalized, not requiring supplemental oxygen; 4, hospitalized, requiring supplemental oxygen; 5, hospitalized, requiring nasal high-flow oxygen therapy, noninvasive mechanical ventilation, or both; 6, hospitalized, requiring ECMO, invasive mechanical ventilation, or both; and 7, death.
† Patients who have turned negative on Day 7 were not recalculated.
ICU= intensive care unit.
There was no death.

### Secondary Outcomes

A total of 15 (51.7%) patients turned viral negative within 7 days after the initiation of the trial (60%, 44% and 50% in the baloxavir, favipiravir, and control group, respectively). One patient in the baloxavir marboxil group, and two patients in the favipiravir group were transferred to ICU within seven days after trial initiation. The patient in baloxavir marboxil group was treated with ECMO ten days after trial initiation. There was no death. Clinical outcomes were showed in Table 2. Together, these results suggest that the addition of either baloxavir or favipiravir under the current dosage to the exiting standard treatment did not provide additional benefits to the clinical outcome in this study.

### Viral Negativity

Throughout the trial, viral load was monitored everyday for each patient. Figure S4 (appendix p7) shows the kinetics of the viral load in the three groups of patients. These results indicate that the addition of either baloxavir or favipiravir didn’t appear to improve the medium T1/2 time for patients to achieve undetectable viral RNA compared to the control group.

### Drug Concentrations in COVID-19 Patients

To determine whether the apparent lack of benefits by the addition of either baloxavir or favipiravir is related to their pharmacological exposure in the COVID-19 patients, drug concentrations were measured in the patients. Overall, 70 plasma samples were collected, 19 samples on Day-1, 28 samples on Day-4, and 23 samples on Day-7. Among these samples, 28 contained baloxavir acid, 20 contained favipiravir, and other samples contained arbidol, lopinavir, ritonavir, and darunavir were detected by UPLC-MS/MS. Administered orally, favipiravir is rapidly absorbed with a t_max_ ranging from 0.5 to 1 hour [6]. Favipiravir total concentration was measured at Day-2 and Day-4 from plasma samples collected before the first favipiravir intake of the day in Ebola-infected patients [6]. As a result, the sampling time was at 0.5–1 hour post dose for the Day-1 samples, and 1–2 hour before dose administration for the Day-4 and Day-7 samples. The plasma concentrations in Day-1, Day-4, and Day-7 samples are listed in Table 3.

**Table 3.** Plasma concentrations of baloxavir and favipiravir on Day-1, Day-4 and Day-7*

|  |  | **Number of patients** | **Observed concentration (µg/mL)†** | **Concentration/EC<sub>50</sub>†** |
| --- | --- | --- | --- | --- |
| Favipiravir | Day 1 | 7 | 32.2 ± 18.7 | ≤2.1 ± 1.2 |
|  | Day 4 | 8 | 11.8 ± 11.6 | ≤0.8 ± 0.7 |
|  | Day 7 | 5 | 21.8 ± 28.7 | ≤1.4 ± 1.8 |
| baloxavir | Day 1 | 9 | 0.016 ± 0.023 | 0.006 ± 0.009 |
|  | Day 4 | 10 | 0.024 ± 0.019 | 0.009 ± 0.007 |
|  | Day 7 | 9 | 0.020 ± 0.013 | 0.008 ± 0.005 |

\*Sampling time: The first day of the sampling time is based on the first administration for favipiravir and baloxavir.
3 Both observed concentration and concentration/EC<sub>50</sub> are presented as mean ± SD.

The total plasma concentration of favipiravir was 11.8 ± 11.6 µg/mL in Day-4 samples, and 21.8 ± 28.7 µg/mL in Day-7 samples (Table 3). With a plasma protein binding of 54%, the free favipiravir trough concentrations on Day-7 was close to the in vitro EC_50_ value of 9.7 µg/mL reported in G.X et al.[2], but lower than the EC_50_ value (>15.7 µg/mL) determined in our study.

Following absorption, baloxavir marboxil is almost entirely converted to its active metabolite, baloxavir acid. The observed concentrations of baloshavir at Day-4 and Day-7 were 0.024 ± 0.019 µg/mL and 0.020 ± 0.013 µg/mL, respectively. The concentrations of baloxavir were much lower than its EC_50_ value of 2.65 µg/mL (5.48 µM), which is consistent with the lack of viral inhibition by this compound.

Arbidol was included in the existing standard treatment of 27 patients in this study, and 57 plasma samples were collected. The plasma concentrations of arbidol were shown in appendix P6 Table S2. With the exception of samples from Patient 15, the plasma concentration in all other samples were lower than the EC_50_ value (1.77 µg/mL).

Lopinavir was included in the existing standard treatment of 21 patients, and the concentration of lopinavir was measured in 36 plasma samples. Appendix P6 Table S2 showed the concentration values of lopinavir. The mean concentrations of lopinavir at Day 1, Day 4, and Day 7 were higher than its EC_50_ (6.54 µg/mL). However, lopinavir is approximately 98 to 99% bound to plasma proteins, so the free drug concentration may not reach the target value. The plasma concentrations of ritonavir and darunavir were shown in Table S2 (appendix P6).

### Adverse Events

Respiratory failure occurred in 14 patients (Table 4). Other adverse events were generally mild or moderate among the three Groups. The most frequent adverse events occurring in the study population were similar among all groups, including elevation of triglyceride (20 events, Figure S5 in appendix p8), liver function abnormality (18 events, Figure S6 in appendix p9), rash (7 events), and diarrhea (4 events) (Table 4). No abnormal serum creatinine was found in all patients.

**Table 4.**
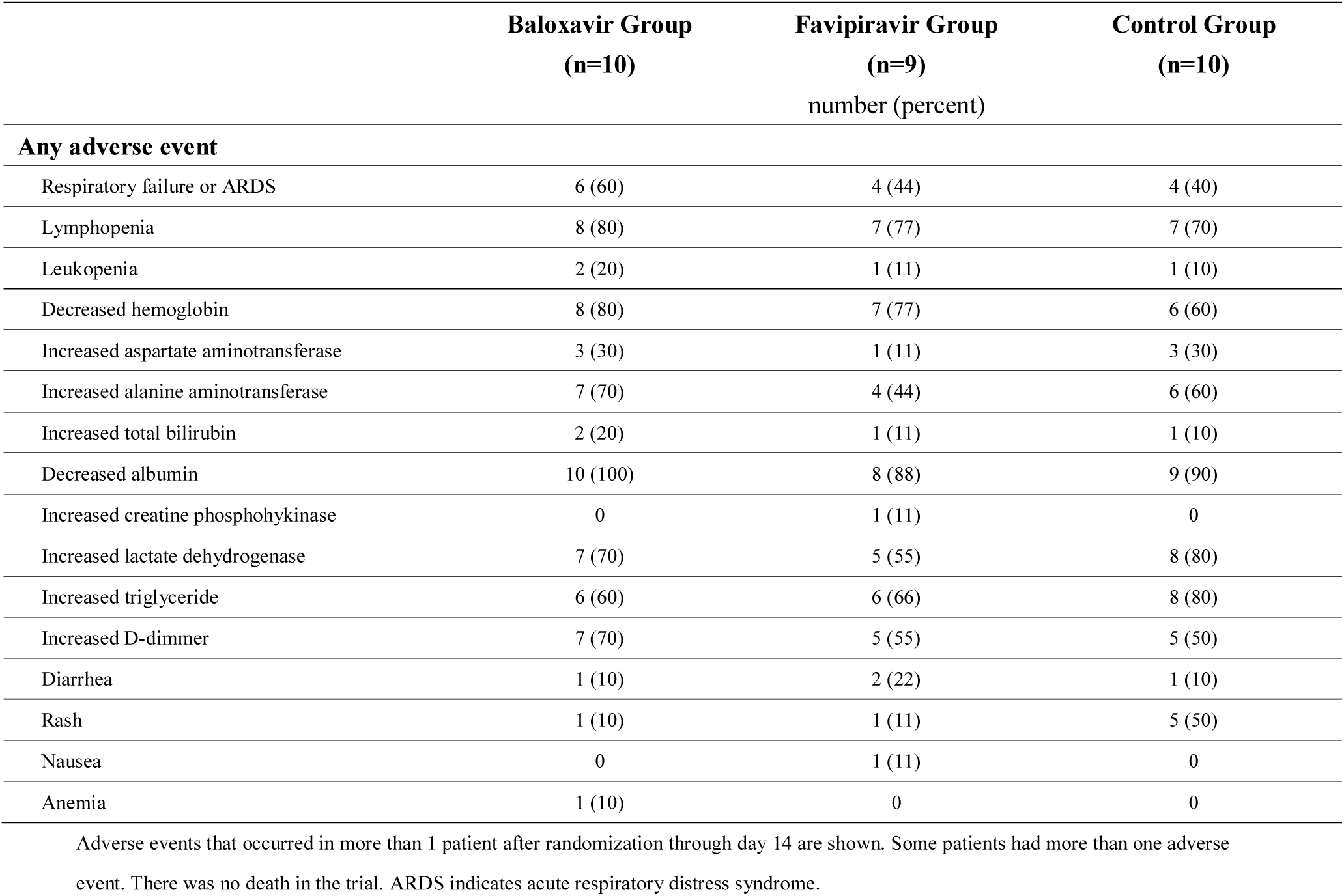
Summary of Adverse Events in the Safety Population

## Discussion

In this study, we assessed the efficacy and safety of baloxavir marboxil and favipiravir in 29 COVID-19 patients who were still virus positive under the current antiviral treatment according to the recommendation of Diagnosis and treatment program of novel coronavirus pneumonia (COVID-19) (Trial Sixth Edition). The results of viral negativity, clinical symptoms, and laboratory tests indicated adding either baloxavir or favipiravir to the current standard treatment did not provide additional benefits to the clinical outcome in this clinical study. Adverse events were generally mild or moderate with no differences in frequency or severity among the three groups. No moderate or severe drug-related unsolicited adverse events were reported during the trial.

Furthermore, in order to provide the pharmacological rationale of drug antiviral activities in vivo, the drug exposure of baloxavir acid, favipiravir, arbidol, lopinavir, ritonavir, and darunavir were also measured. The drug concentrations of favipiravir in our study were slightly lower than that of Ebola patients[6], which may be caused by different dosage. The exposure of baloxavir and lopinavir/ritonavir in these COVID-19 patients were similar as that in influenza[7] or HIV-infected patients [8] which reported in the previous studies. However, the pre-dose drug concentrations of abidol in this study were 2–3 times higher than the steady-state concentrations of C_min,ss_ (0.17 µg/mL) in healthy Chinese volunteers[9], which may be due to the drug-drug interaction between abidol and ritonavir or cobicistat based on CYP3A4[10]. Similarly, comparing the minimum plasma concentration (C_min_,1.3 µg/mL) of darunavir in HIV-1-infected adults [11], the plasma concentration in this study were much higher. Although, the results from this trial showed that the free drug concentrations of these five antiviral drugs were generally lower than their respective EC_50_ values. The latest published clinical trial report that patients with severe COVID-19 had no benefit from lopinavir-ritonavir treatment[12], which to some extent confirms our view.

Our findings do not support adding either baloxavir marboxil or favipiravir under the current dosage as antiviral agents to the existing standard treatment in COVID-19 patients. However, this conclusion should be taken with caution for several reasons. In this study, the analysis relied only on plasma concentrations and in vitro antiviral activity against SARS-CoV-2, while intracellular concentrations of the active phosphorylated moiety were not available. For favipiravir, the intracellular concentrations of the active metabolite has been shown to be associated with antiviral efficacy, instead of the plasma concentrations of the parent molecule [13]. Secondly, the poor correlation could be due to the delay between infection and treatment initiation. Viral dynamic modelling shows that a drug affecting viral replication, will only have a limited impact on viraemia if treatment is initiated after the viraemia peak, regardless of drug efficacy[14]. Since the subjects of this trial were those who were still positive after the treatment of the recommended scheme, the optimum time to start antiviral therapy may have been missed. There are many reasons for the failure of viral load reduction, some of which are related to the pharmacology of drugs. We are not sure if these drugs will appear in respiratory secretions, or if they are not so effective against SARS-CoV-2.

There are some limitations to the current study. Firstly, the subjects were all under treatment with other medication. The treatment scheme and medication time before the initiation of the trial were different among the patients, which makes their progression of the disease at the beginning of the trial quite different. There was therefore a risk of influencing the results and conclusions. During a rapidly evolving COVID-19 situation, it was difficult to obtain large number of newly detected cases without previous treatment. Second, patients in favipiravir group showed oldest average age and shortest time from symptom onset to randomization, even though, the clinical performance of favipiravir group was not inferior to the other two groups. However, we are not sure if the efficacy of favipiravir under current dosage is underestimated because its drug exposure does not reach the EC_50_ value. Third, the relatively small sample size of our study poses an additional limitation. Nevertheless, it was conclusive that the free plasma concentrations of these antiviral drugs did not reach their respective EC_50_ values, which can be almost certainly that the drugs have no effect against SARS-CoV-2 at the dose as mentioned above. Our research cannot be simply regarded as negative results, as it is very meaningful for clinical treatment of COVID-19 in global outbreak. In addition, our exploratory research provides useful information for further studies to find the best strategy for application of these drugs.

## Conclusion

This exploratory trial does not prove that baloxavir marboxil was effective in COVID-19 patients. Because the free drug concentration of baloxavir marboxil is far below than its EC_50_ values (more than 100 times). Under the current dosage, the insufficient exposure of favipiravir also resulted in no additional antiviral benefit by adding favipiravir to the existing standard treatment. Administration of favipiravir at different dosage or at different stages of COVID-19 (for example, early stage) deserves further study. Additional studies are needed to confirm the no clinical benefit from the current standard treatment drugs.

## Data Availability

The complete de-identified patient dataset, statistical plan, and consent forms will be made available on request to YQ Q 1 year after publication, with no time limit.

## Supplementary Data

Supplementary materials are available online. Consisting of data provided by the authors to benefit the reader, the posted materials are not copyedited and are the sole responsibility of the authors, so questions or comments should be addressed to the corresponding author.

## Acknowledgments

This article was supported by Zhejiang Provincial Science and technology department key R & D plan emergency project (No. 2020c03123–8). We acknowledge Dr. Chengyong Yang and Dr. Guofeng Cheng for their technical support to this study. Dr. Yang and Dr. Cheng were given the opportunity to review the manuscript for medical and scientific accuracy.

## Reference

1. Chan JF, Yuan S, Kok KH, To KK, Chu H, Yang J, Xing F, Liu J, Yip CC, Poon RW, Tsoi HW, Lo SK, Chan KH, Poon VK, Chan WM, Ip JD, Cai JP, Cheng VC, Chen H, Hui CK, Yuen KY. A familial cluster of pneumonia associated with the 2019 novel coronavirus indicating person-to-person transmission: a study of a family cluster. Lancet 2020;395:514–23.

2. Wang M, Cao R, Zhang L, Yang X, Liu J, Xu M, Shi Z, Hu Z, Zhong W, Xiao G. Remdesivir and chloroquine effectively inhibit the recently emerged novel coronavirus (2019-nCoV) in vitro. Cell Res 2020;30:269–71.

3. Hayden FG, Sugaya N, Hirotsu N, Lee N, de Jong MD, Hurt AC, Ishida T, Sekino H, Yamada K, Portsmouth S, Kawaguchi K, Shishido T, Arai M, Tsuchiya K, Uehara T, Watanabe A; Baloxavir Marboxil Investigators Group. Baloxavir Marboxil for Uncomplicated Influenza in Adults and Adolescents. N Engl J Med 2018;379:913–23.

4. Hayden FG, Shindo N. Influenza virus polymerase inhibitors in clinical development. Curr Opin Infect Dis 2019;32:176–86.

5. Abdelnabi R, Morais ATS, Leyssen P, Imbert I, Beaucourt S, Blanc H, Froeyen M, Vignuzzi M, Canard B, Neyts J, Delang L. Understanding the Mechanism of the Broad-Spectrum Antiviral Activity of Favipiravir (T-705): Key Role of the F1 Motif of the Viral Polymerase. J Virol 2017;91: pii: e00487–17.

6. Nguyen TH, Guedj J, Anglaret X, Laouenan C, Madelain V, Taburet AM, Baize S, Sissoko D, Pastorino B, Rodallec A, Piorkowski G, Carazo S, Conde MN, Gala JL, Bore JA, Carbonnelle C, Jacquot F Raoul H, Malvy D, de Lamballerie X, Mentré F, JIKI study group. Favipiravir pharmacokinetics in Ebola-Infected patients of the JIKI trial reveals concentrations lower than targeted. PLoS Negl Trop Dis 2017;11:e0005389.

7. Abraham GM, Morton JB, Saravolatz LD. Baloxavir: A Novel Antiviral Agent in the Treatment of Influenza. Clin Infect Dis 2020: pii: ciaa107.

8. Molto J, Valle M, Blanco A, Negredo E, DelaVarga M, Miranda C, Miranda J, Domingo P, Vilaró J, Tural C, Costa J, Barbanoj MJ, Clotet B. Lopinavir/ritonavir pharmacokinetics in HIV and hepatitis C virus co-infected patients without liver function impairment: influence of liver fibrosis. Clin Pharmacokinet 2007;46:85–92.

9. Sun Y, He X, Qiu F, Zhu X, Zhao M, Li-Ling J, Su X, Zhao L. Pharmacokinetics of single and multiple oral doses of arbidol in healthy Chinese volunteers. Int J Clin Pharmacol Ther 2013;51:423–32.

10. Deng P, Zhong D, Yu K, Zhang Y, Wang T, Chen X. Pharmacokinetics, metabolism, and excretion of the antiviral drug arbidol in humans. Antimicrob Agents Chemother 2013;57:1743–55.

11. Tashima K, Crofoot G, Tomaka FL, Kakuda TN, Brochot A, Van de Casteele T, Opsomer M, Garner W, Margot N, Custodio JM, Fordyce MW, Szwarcberg J. Cobicistat-boosted darunavir in HIV-1-infected adults: week 48 results of a Phase IIIb, open-label single-arm trial. AIDS Res Ther 2014;11:39.

12. Cao B, Wang Y, Wen D, Liu W, Wang J, Fan G, Ruan L, Song B, Cai Y, Wei M, Li X, Xia J, Chen N, Xiang J, Yu T, Bai T, Xie X, Zhang L, Li C, Yuan Y, Chen H, Li H, Huang H, Tu S, Gong F, Liu Y, Wei Y, Dong C, Zhou F, Gu X, Xu J, Liu Z, Zhang Y, Li H, Shang L, Wang K, Li K, Zhou X, Dong X, Qu Z, Lu S, Hu X, Ruan S, Luo S, Wu J, Peng L, Cheng F, Pan L, Zou J, Jia C, Wang J, Liu X, Wang S, Wu X, Ge Q, He J, Zhan H, Qiu F, Guo L, Huang C, Jaki T, Hayden FG, Horby PW, Zhang D, Wang C. A Trial of Lopinavir-Ritonavir in Adults Hospitalized with Severe Covid-19. N Engl J Med 2020; [Epub ahead of print].

13. Bazzoli C, Jullien V, Le Tiec C, Rey E, Mentre F, Taburet AM. Intracellular Pharmacokinetics of Antiretroviral Drugs in HIV-Infected Patients, and their Correlation with Drug Action. Clin Pharmacokinet 2010;49:17–45.

14. Madelain V, Oestereich L, Graw F, Nguyen TH, de Lamballerie X, Mentre F, Günther S, Guedj J. Ebola virus dynamics in mice treated with favipiravir. Antiviral Res 2015;123:70–7.

